# Understanding mental health trends during COVID-19 pandemic in the United States using network analysis

**DOI:** 10.1101/2022.10.10.22280933

**Authors:** Hiroko Kobayashi, Raul Saenz-Escarcega, Alexander Fulk, Folashade B. Agusto

## Abstract

The emergence of COVID-19 in the United States resulted in a series of federal and state-level lock-downs and COVID-19 related health mandates to manage the spread of the virus. These policies may negatively impact the mental health state of the population. This study focused on the trends in mental health indicators following the COVID-19 pandemic amongst four United States geographical regions, and political party preferences. Indicators of interest included feeling anxious, feeling depressed, and worried about finances. Survey data from Delphi Group in Carnegie Mellon University were analyzed using clustering algorithms and dynamic connectome obtained from sliding window analysis. United States maps were generated to observe spatial trends and identify communities with similar mental health and COVID-19 trends. Between March 3rd, 2021 and January 10th, 2022, states in the south geographic region showed similar trends for reported values of feeling anxious and worried about finances. There were no identifiable communities resembling geographical regions or political party affiliation for the feeling depressed indicator. We observed a high degree of correlation among southern states as well as within republican states, where the highest correlation values from the dynamic connectome analysis for feeling anxious and feeling depressed variables seemingly overlapped with an increase in COVID-19 related cases, deaths, hospitalizations, and rapid spread of the COVID-19 Delta variant.

## 1 Introduction

Following an initial outbreak in 2019 in Wuhan, China, the novel coronavirus disease (COVID-19) spread rapidly across the world, resulting in nearly 4.2 million infected and approximately 85 thousand dead within the first year of being identified in 2020 [33]. By March 22 of 2020, the World Health Organization (WHO) declared COVID-19 a global pandemic [8]. This declaration resulted in several countries adopting various protocols to collect data from local counties and municipalities to make informed decisions on COVID-19 policies to curb the spread of COVID-19 in their regions [13, 3]. However, due to limited information available on the novel disease, many countries opted to implement large-scale control measures such as lockdowns that had far-reaching impacts on the general population [13].

To manage the spread of COVID-19 in the United States, the federal government and state governments implemented a variety of health mandates [14]. However, the federal government at this time opted for a hands-off approach to COVID-19 and allowed individual states to decide on the best ways to limit the spread of this deadly disease [1]. This variation in leadership led to differing outcomes in terms of the spread of COVID-19 [26]. These mandates to protect the public ranged from school and work closures to stay-at-home orders, all of which can have important effects on several aspects of an individual’s life. In addition, the increasing incidence of COVID-19 and COVID-19 related deaths likely increased individual’s pandemic related worry [12]. These potential stressors have, and continue to impact each member of the population to varying degrees. Policies were enacted to enforce many decisions related to the pandemic and may have adverse mental health effects including feelings of anxiety and depression. Elevated adverse mental health conditions were reported at a higher disproportionate rate in the second quarter of 2020 (25.5%) compare to the last quarter of 2019 (8.1%). [9, 21]. Several countries, including the United States, have input measures to allow for reporting of psychological distress among the population via surveys to better tailor support and resources for their respective populations [31, 32].

Several studies have concluded that lockdowns and other policies related to COVID-19 can increase mental health burden, especially for vulnerable groups [2, 18, 29]. Certain policies related to COVID-19, such as government funds being issued to the general population via stimulus checks, also have the potential for positive impacts on both physical and mental health [11]. Vaccines also likely decreased the prevalence of mental health issues since their initial roll-out in late 2020 [23]. Though a potentially large portion of the public have displayed hesitance towards vaccination and thus may be experiencing similar levels of mental distress [22]. Due to the various possible mental health outcomes that arise from each of these scenarios, it is important to evaluate data in a manner that facilitates dynamic interpretation across time in a clear and concise way.

For instance, Amico and Bulai [4] used network analysis to explore the interactions of COVID-19 among the various regions of Italy and the impact of Italy governmental policies in response to the spread of SARS-Cov-2. To analyze the network interaction between regions in Italy, they used six indicators (namely the number of hospitalized individuals in IC, number of hospitalized individuals with symptoms, number of individuals in home isolation, number of new positive, number of discharged healed, and number of deceased individuals) to form a correlation network refer as “Covidome”. Their results showed that there was a distinct north-south clustering of the regions in the country. Furthermore, they found distinct difference of the Covidome fluctuations between the first and second wave of the pandemic based on region-specific political choices.

In this study, we apply network and clustering analysis to understand the connectivity between states and how COVID-19 impacts mental health across the United States using COVID-19 related mental health indicators such as feeling anxious, feeling depressed, and worried about finances. We follow the approach in Amico and Bulai [4] and study the covariance matrix of the mental health indicators (which we refer to as the “mental health functional connectome”). We then compare the results from this study with results from Fulk et al [10] that investigated the effect of several COVID-19 related indicators variables on anxiety and depression in the general population using the same data during the same time period as this study. The rest of the paper is organized as follows: Section 2 gives an overview of the methods used in the analyses, including a description of the data; Section 3 describes the results of the study. Section 4 presents the discussion, where we put our results into the context of policies that were implemented and also evaluate how our results compares to Fulk et al [10] analysis of this data, as well as their conclusion.

## 2 Methods

### 2.1 Mental health related data and policy timeline

#### Mental health data

This research is based on survey results from Carnegie Mellon University’s Delphi Group. The Carnegie Mellon University U.S. COVID-19 Trends and Impact Survey (Delphi US CTIS [24]) was distributed in partnership with Facebook in the form of a daily voluntary survey that Facebook users could respond to. Survey questions ranged from those concerning physical health, the economic impact of COVID-19, to mental health and behavioural prompts. Participant responses were collected, aggregated, and made publicly available. For this study, three indicators of interest were selected to represent the impact of COVID-19 on mental health. These three indicators were (i) the percentage of participants who experienced feelings of anxiety within the last 7 days, (ii) the percentage of participants who felt worried about their finances for the following month, and (iii) the percentage of participants who felt depressed within the last 7 days. We refer to these three indicators as feeling anxious, feeling depressed, and worried about finances throughout this paper. Daily confirmed COVID-19 cases and daily deaths from the Center for Systems Science and Engineering at Johns Hopkins University [6] were also included in this study. These indicators were used to assess whether there was any relationship between trends in cases or deaths and the trends in mental health indicators.

The survey results collected can be categorized in two separate time-frames, the first starting from September 8th, 2020 running until March 2nd, 2021, and the second beginning from March 2nd, 2021 to January 10th, 2022. This separation is due to a change in the survey questions asked about anxiety and depression; in the first timeframe, the survey asked participants whether they experienced feelings of anxiety or depression for the past 5 days. From March 2nd, 2021, the survey asked whether the participants experienced feelings of anxiety or depression for the past 7 days, resulting in data collected from March 2nd differing from that of earlier dates. Data analysis was only conducted using data collected from the second time-frame due to missing data points for mental health indicators from the first time-frame.

Daily COVID-19 related cases, deaths, and hospitalizations between March 2nd, 2021 to January 10th, 2022 were plotted to identify waves in the spread of COVID-19. Three COVID-19 waves were identified, as can be seen in Figure 1. These waves are most distinguishable in Fig 1 (c), depicting daily COVID-19 related hospitalizations across four geographical regions in the US. Based on these waves, we split the data into three periods, reflective of the start and end of each wave. The dates for each period are (a) April 1st, 2021 to July 1st, 2021, (b) July 2nd, 2021 to November 11th, 2021, and (c) November 2nd, 2021 to January 10th, 2022.

**Figure 1:**
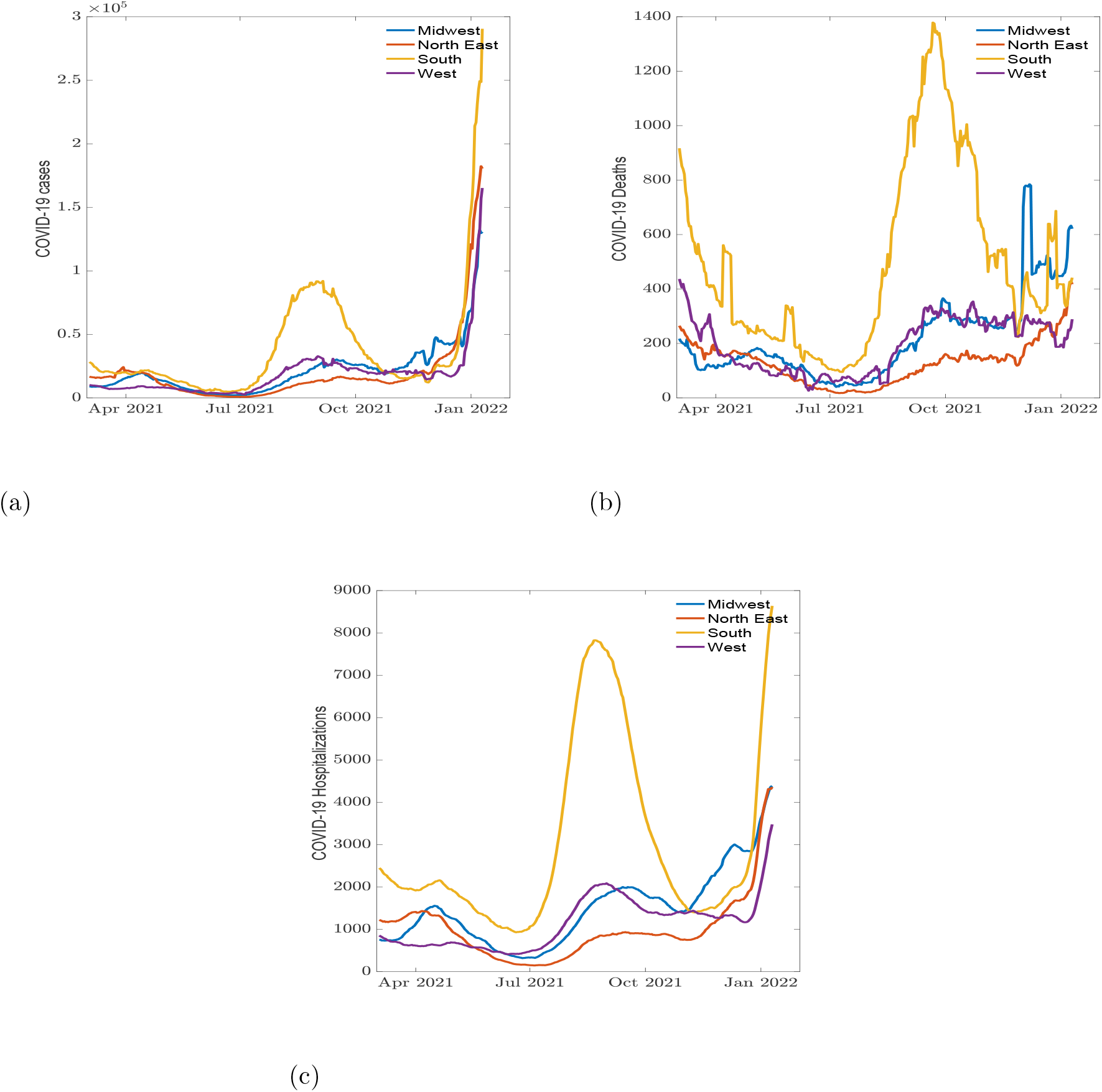
Daily COVID-19 (a) cases, (b) reported deaths, and (c) hospitalization cases across demographic regions.

In addition to looking at individual states, four United States geographical regions (midwest, northeast, south, and west) as well as political party preference (Democratic, Republican) were used to split the states to see if any mental health trends among states clustered similarly to these geographically or politically established communities. There is variation in how the country can be split geographically; in many cases it is based on the historical relationship certain regions have among one another. These relationships can stem from a similar religious community, cultural similarities, shared historical significance or similar climates in the region [20]. In this study, we referenced the United States Census Bureau geographic map of the country which is compromised of four regions: midwest, northeast, south and west [17]. The political party preference was determined from the results of the 2020 presidential election, senate, and overall state party registration percentage [30]. The regions and political party each state belongs to can be seen in Figure 2.

**Figure 2:**
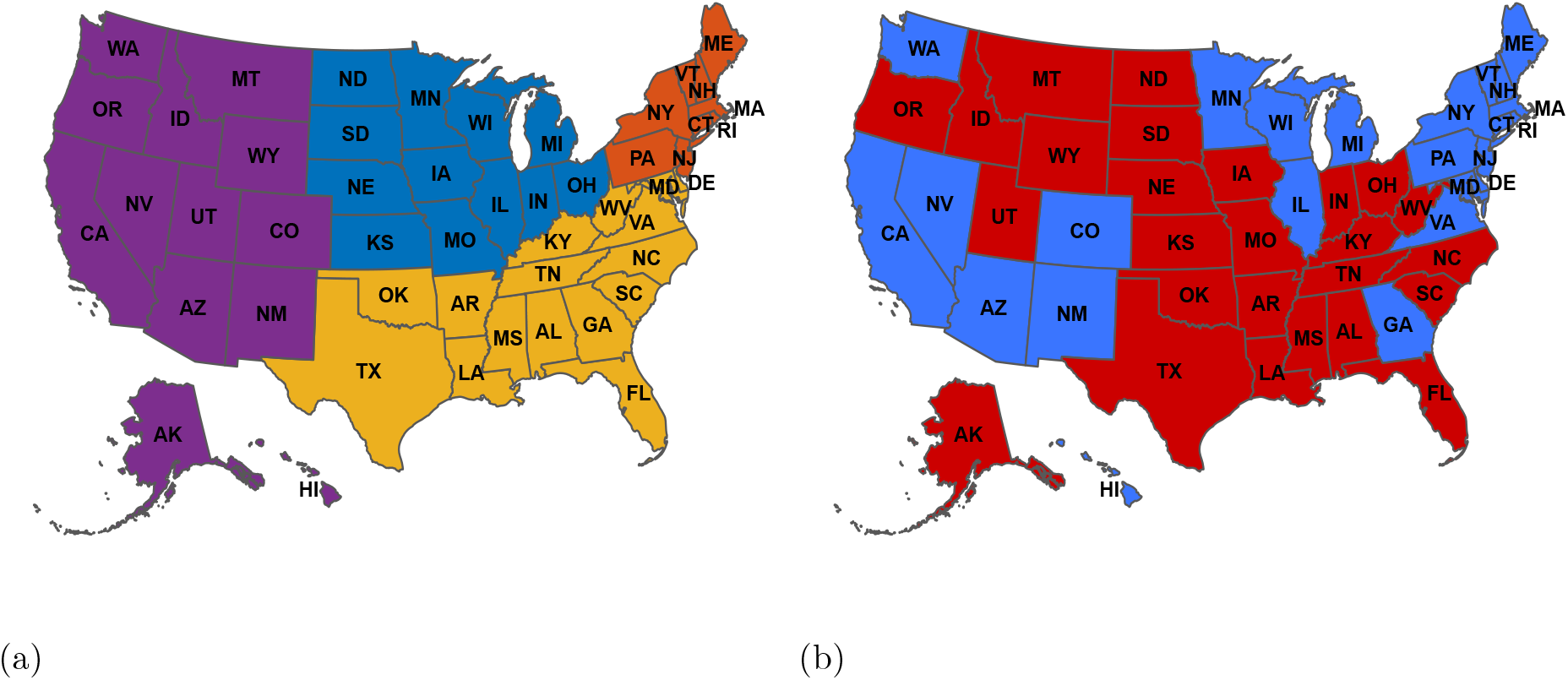
Regional and political maps of the United States. (a) A regional map where blue represents the midwest, orange represents the northeast, yellow represents the south, and purple represents the west; (b) Political map where blue represents Democratic, and red represents Republican party preference. The abbreviations are those given in the International society for organization standard for the United States (ISO 3166-2, website: https://www.iso.org/obp/ui/#iso:code:3166:US)

Similar to Figure 1, the daily COVID-19 related cases, deaths, and hospitalizations were grouped based on each state’s political preference, and were plotted to identify waves in the spread of COVID-19 (see Figure 10 in Appendix A). The survey results on the mental health indicators were also plotted based on geographical regions (see Figure (3)) as well as political preference (see Figure 11 in Appendix B).

#### COVID-19 policy timeline

Next we list a timeline of a few policies implemented during the COVID-19 outbreak in the country. Some policies listed in this timeline reflect efforts implemented to tackle the outbreak, such as nation-wide vaccine roll out for all adults, unemployment benefits, and an eviction moratorium. Depending on each policy’s target population and topic, as well as how we hypothesized these policies would impact individuals’ mental health, the policies were categorized as related to anxiety and depression, or worried about one’s finances.

- March 11, 2021 – Biden signs a coronavirus relief package bill including extended unemployment benefits and a third stimulus check^2^
- April 19, 2021 – all adult Americans eligible for vaccines by 4.19^3^
- April 21, 2021 – Tax credit for small businesses and non-profits, covers 80h of leave, $511/day^2^
- May 13, 2021 – CDC guidance on masking updated, no longer recommends masks indoors or outdoors for fully vaccinated people regardless of number of people gathered^3^
- July 3, 2021 – CDC states more than 50% of cases are Delta variant^3^
- July 31, 2021 – Moratorium on eviction expires^2^.
- August 3, 2021 – CDC issues eviction moratorium specifically in areas with ‘substantial or high’ rates of COVID transmission^2^
- August 26, 2021 – Moratorium ends by Supreme Court ruling^2^
- September 6, 2021 – Unemployment benefits end for many individuals^2^
- October 20, 2021 – Biden announces plan for distributing vaccine in children 5
- 11 if the vaccine is authorized, FDA authorizes Moderna and J&J vaccine in eligible populations, and allows mixing vaccine doses (can use different primary and booster shots)^3^
- December 9, 2021 – FDA expands Pfizer-BioNTech booster eligibility to 16- and 17-year olds^3^
- December 27, 2021 – CDC shortens isolation and quarantine period ^3^

### 2.2 Clustering and correlation networks

For each indicator, we obtained a Pearson’s correlation coefficient matrix, 𝕡 [16]. The matrix entry 𝕡_*i,j*_ is obtained by calculating the Pearson’s correlation coefficient from state *i*’s and state *j*’s time series data, where *i* and *j* may refer to any state in the United States. Each Pearson’s correlation coefficient matrix was used as an adjacency matrix, which defines the connectivity of states (with respect to a particular indicator) in a network. If 𝕡_*i,j*_ ≠ 0, then states *i* and *j* are connected. Otherwise, they are disconnected. Moreover, 𝕡_*i,j*_ *>* 0 (positive connection) indicates a positive correlation, while 𝕡_*i,j*_ *<* 0 (negative connection) indicates a negative correlation; the more positive (or negative) 𝕡_*i,j*_ is, the stronger the positive (or negative) correlation [16].

A cluster consists of states (or nodes) that are more similar (with respect to a particular indicator) to each other than to states in other clusters. Whether a given pair of states are grouped together depends on both how they are connected to each other and how they are connected to other states in the network. We utilized Traag’s implementation of the Leiden algorithm [25] in Python 3.9.7 from Spyder version 5.2.2 (specifically, the find partition and optimise partition multiplex functions from the leidenalg package [28]). Briefly, these functions were used as follows: Each network was split into a positive and negative sub-network. The positive sub-network contained only the positive connections from the original network, and the negative sub-network contained only the negative connections from the original network [28]. We applied the find partition function to both sub-networks to obtain clusters for each, and we thereafter applied the optimise partition multiplex function to both sets of clusters to obtain a single set of clusters for the original network.

The clusters returned by Leiden’s algorithm are not unique, meaning that if Leiden’s algorithm is run multiple times, the clusters may not be the same. Consensus clustering addresses this issue by grouping states based on multiple runs of the Leiden algorithm [19]. Specifically, the Leiden algorithm is ran multiple times with a given network (hereafter referred to as the original network), and after all runs are completed, the frequencies at which states are grouped together are used to create a new adjacency matrix (and thus a new network); the Leiden algorithm is then ran with this new network. The resulting clusters are referred to as consensus clusters because they represent the “consensus” of multiple Leiden algorithm runs with the original network. The number of runs necessary may be assessed by tracking how the above-mentioned frequencies change as more runs are performed. We manually implemented consensus clustering in Python with 300 runs of the Leiden algorithm [28]. We verified that 300 runs were sufficient by visually inspecting how frequencies changed with more runs.

Once these matrices were collected, the Louvain algorithm was used in an iterative process called consensus clustering to determine the best community membership for each state pair. Consensus clustering runs the Louvain algorithm multiple times to assess which states go together (often showing similar indicator trends) most frequently and consistently and assigns memberships or communities to each state. These memberships were stored in consensus matrices. Each run consisted of 300 iterations, and a minimum of 200 extension runs were conducted. Subsample size was 90% (45 states) to introduce variation, and further extensions were applied following visual analysis of state membership stabilization using progress plots that showed the modularity score change with each iteration. The results from the final iteration for each indicator consensus matrix were stored, then mapped onto United States maps to visually assess the memberships for each variable. In addition, the final iteration results were also used to construct an allegiance matrix. The allegiance matrix gives “the probability for two regions of being in the same community across all Covid indicators” [4, 7].

### 2.3 Dynamic connectome analysis

We follow the approach by Amico and Bulai [4], and use the sliding window analysis to understand the link between mental health indicators, government policy and their relationship across geographic regions and political parties. The sliding window analysis is a technique constructed by taking a fixed time interval to explore the different indicators at various times frames. This technique is commonly used in the field of neuroscience to examine brain network dynamics [4,5]. In our sliding window analysis, we take a fixed window of 30 days, and slide this by 1 day to examine the network dynamics of our mental health indicators. Each data point is thus a correlation value taken across the 30 days within a window, which can be plotted to display the dynamic fluctuations in the variable of interest across the time-frame used. We evaluated the differences between each region according to four different areas (midwest, northeast, south, west) and their respective political affiliation (see Figure 2).

## 3 Results

All analyses were conducted in R 2021 version, Python 3.9.7 from Spyder version 5.2.2 and figures in MATLAB R2021b. The Delphi US CTIS data was accessible through the Covidcast library.

### Consensus and allegiance clustering

Figure 4 shows the membership results of the consensus clustering for the three mental health indicators. The membership results are shown as distinct clusters separated by color on the United States maps in Figure 2 (a). Figures 4 (a) and Figure 4 (c) show similar trends, with no clear community distinction other than a slight South region clustering (darker blue). The south region clustering observed in both Figures 4(a) and Figure 4 (c) looks like the map of the republican party affiliation shown in Figure 2 (b). Figure 4 (b) shows no distinct clustering pattern, indicating that the communities with similar trends for the feeling depressed indicator did not cluster in manners reflecting geographical regions or political party affiliation shown in Figure 2.

The membership results across the three mental health indicators were taken to construct an allegiance matrix (See Section 2.2 for detailed information). The map on Figure 5 (a) and the corresponding heat-map of the allegiance matrix is shown in Figure 5 (b). These indicate which states were most likely to be grouped together (and exhibit similar trends) across all three mental health indicators. The map shows three main clusters, of which the light blue colored states are most interesting. The clustering of the light blue states closely looks like the south geographic region, with the exception of Virginia, West Virginia, and Arkansas, which were not as frequently grouped together with the rest of the south. The light blue colored states also include California, North Carolina, and Nevada, which are non-southern states.

### Dynamic connectome

Next, we consider the results of the dynamic connectome obtained by taking snapshots of the connectome in overlapping time windows of 30 days using sliding window analysis. The dynamic connectome results include changes in both correlation values (see Figures 6 and 8) as well as eigenvector centrality values (see Figures 7 and 9) for the mental health indicators. Analyses were conducted separately for demographic regions and political party preference. As mentioned in Section 2.1, the survey data collected was split into 3 periods corresponding to the three waves seen within the time-frame used. Maximum and minimum correlation values within each period were recorded, as seen in Tables 1 and 3. The eigenvector centrality values (Tables 2 and 4) work to verify the results of the correlation values; these values highlight regions or parties which are most important, especially in times of lower correlation. To minimize confusion when referencing the dynamic connectome results of each of the three indicators, we denote all future references by use of the term variable rather than indicator. Changes in the collected survey data will be discussed in the results and discussion as indicators, while discussing the results of dynamic connectome on this data (such as correlation values and eigenvector centrality value changes) will be marked by the word variable.

**Table 1:** Minimum and maximum values of correlation time series for feeling anxious, feeling depressed, worried about finances between demographic regions

| Regions | Wave | Anxious<br>Min | Anxious<br>Max | Depressed<br>Min | Depressed<br>Max | Finance<br>Min | Finance<br>Max |
| --- | --- | --- | --- | --- | --- | --- | --- |
| Midwest | 1 | -0.0171 | 0.4514 | -0.0690 | 0.5958 | -0.0565 | 0.8554 |
| Midwest | 2 | -0.0317 | 0.4755 | -0.0689 | 0.4213 | -0.0668 | 0.1232 |
| Midwest | 3 | -0.0426 | 0.0836 | -0.0291 | 0.0614 | -0.0186 | 0.1428 |
| North East | 1 | -0.0648 | 0.4523 | -0.0033 | 0.5166 | -0.0524 | 0.6376 |
| North East | 2 | -0.0762 | 0.4413 | -0.0867 | 0.3642 | -0.1000 | 0.1245 |
| North East | 3 | -0.0677 | 0.0478 | -0.0330 | 0.1222 | -0.0688 | 0.2052 |
| South | 1 | -0.0421 | 0.4498 | -0.0525 | 0.3267 | -0.0216 | 0.8694 |
| South | 2 | -0.0365 | 0.5753 | -0.0453 | 0.5160 | 0.0026 | 0.4765 |
| South | 3 | -0.0382 | 0.0609 | -0.0464 | 0.0583 | -0.0301 | 0.1457 |
| West | 1 | -0.0212 | 0.2302 | -0.0524 | 0.3317 | -0.0396 | 0.7274 |
| West | 2 | -0.0559 | 0.3675 | -0.0542 | 0.3576 | -0.0472 | 0.1770 |
| West | 3 | -0.0694 | 0.0633 | -0.0633 | 0.0291 | -0.0143 | 0.2447 |

**Table 2:** Minimum and maximum values of eigenvector centrality time series for feeling anxious, feeling depressed, worried about finances between demographic regions

| Regions | Wave | Anxious | Anxious | Depressed | Depressed | Finance | Finance |
| --- | --- | --- | --- | --- | --- | --- | --- |
|  |  | Min | Max | Min | Max | Min | Max |
| Midwest | 1 | 0.1014 | 0.7760 | 0.1446 | 0.8178 | 0.1654 | 0.9493 |
| Midwest | 2 | 0.2104 | 0.7737 | 0.1273 | 0.7464 | 0.2076 | 0.4684 |
| Midwest | 3 | 0.1345 | 0.4932 | 0.2303 | 0.5749 | 0.1148 | 0.5763 |
| North East | 1 | 0.1111 | 0.7954 | 0.3198 | 0.8142 | 0.2076 | 0.8642 |
| North East | 2 | 0.1128 | 0.7650 | 0.1445 | 0.7820 | 0.1613 | 0.6512 |
| North East | 3 | 0.1557 | 0.5786 | 0.1622 | 0.6298 | 0.1871 | 0.5992 |
| South | 1 | 0.1966 | 0.7666 | 0.1523 | 0.6981 | 0.1617 | 0.9465 |
| South | 2 | 0.1141 | 0.8252 | 0.2042 | 0.7711 | 0.0791 | 0.7831 |
| South | 3 | 0.1344 | 0.4915 | 0.1982 | 0.4606 | 0.1799 | 0.5155 |
| West | 1 | 0.1340 | 0.6420 | 0.1863 | 0.7038 | 0.1804 | 0.8739 |
| West | 2 | 0.1778 | 0.7232 | 0.2450 | 0.6793 | 0.1360 | 0.5625 |
| West | 3 | 0.2715 | 0.4480 | 0.1966 | 0.4882 | 0.1225 | 0.6512 |

**Table 3:** Minimum and maximum values of correlation time series for feeling anxious, feeling depressed, worried about finances between political parties

| Parties | Wave | Anxious<br>Min | Anxious<br>Max | Depressed<br>Min | Depressed<br>Max | Finance<br>Min | Finance<br>Max |
| --- | --- | --- | --- | --- | --- | --- | --- |
| Democratic | 1 | -0.0196 | 0.3994 | -0.0151 | 0.3365 | -0.0061 | 0.7423 |
| Democratic | 2 | -0.0316 | 0.4007 | -0.0217 | 0.3284 | -0.0081 | 0.1117 |
| Democratic | 3 | -0.0111 | 0.0991 | -0.0258 | 0.0663 | -0.0077 | 0.0770 |
| Republican | 1 | -0.0145 | 0.2358 | -0.0277 | 0.2810 | -0.0199 | 0.8104 |
| Republican | 2 | -0.0196 | 0.4394 | -0.0293 | 0.4507 | -0.0085 | 0.1735 |
| Republican | 3 | -0.0181 | 0.0787 | -0.0277 | 0.0301 | 0.0077 | 0.1764 |

**Table 4:** Minimum and maximum values of eigenvalue centrality time series for feeling anxious, feeling depressed, and worried about finances between political parties

| Parties | Range | Anxious | Anxious | Depressed | Depressed | Finance | Finance |
| --- | --- | --- | --- | --- | --- | --- | --- |
|  |  | Min | Max | Min | Max | Min | Max |
| Democratic | 1 | 0.1539 | 0.6633 | 0.2133 | 0.6597 | 0.2062 | 0.8785 |
| Democratic | 2 | 0.1781 | 0.6905 | 0.1651 | 0.6492 | 0.1448 | 0.5127 |
| Democratic | 3 | 0.1810 | 0.4663 | 0.1176 | 0.4621 | 0.1585 | 0.4794 |
| Republican | 1 | 0.0804 | 0.5932 | 0.1624 | 0.6461 | 0.1684 | 0.9139 |
| Republican | 2 | 0.1764 | 0.7153 | 0.1173 | 0.7237 | 0.1437 | 0.5652 |
| Republican | 3 | 0.2116 | 0.4143 | 0.1931 | 0.3836 | 0.1490 | 0.5483 |

### Regional dynamic connectome

For the regional dynamic connectome results of the feeling anxious variable, both the lowest minimum (−0.0648) and highest maximum (0.4523) correlation values within the first time period were observed from the northeastern region (see Figure 6 (a) and Table 1). In the second period, the minimum correlation value (−0.0762) remained in the northeast, and the maximum correlation value (0.5753) was recorded from the south. The final period showed the minimum correlation value (−0.0694) in the West, and the maximum correlation was seen in the midwest region (0.0836).

For the correlation values in the feeling depressed variable, the first range had both maximum (0.5958) and minimum (−0.0690) correlation values in the midwest region (see Figure 6 (b) and Table 1). The second period exhibited a similar pattern to the feeling anxious variable, with the northeast having the minimum correlation (−0.0867) and the south with the maximum correlation (0.5160). The final period continued with the west having the minimum correlation (−0.0633) among all four regions, and the northeast with the maximum correlation (0.1222). The pattern of the plot of both the feeling anxious and feeling depressed variable were similar, where the graph initially shows a sharp negative slope, but has a large peak during the second period.

The remaining variable, worried about finances in Figure 6 (c) and Table 1, exhibited higher maximum correlation values in the first time period, which decreased with each coming period. During the first period, the midwest had the minimum correlation value (−0.0565), and the south had the maximum correlation value (0.8694). The south region maintained the highest correlation values (0.4765) during the second period, and the minimum value (−0.1000) was in the northeast. For the third period, the northeast still had the lowest correlation value (−0.0688) within the four regions, and the west had the maximum value (0.2447).

For all three variables across each period, the recorded minimum values were very weak negative correlations (all under -0.1), and the states could be considered to have no correlation within each region. The third period maximum values also all have weak positive correlation values (between 0.0291 to 0.2447) compared to the first and second period where most values are considered moderate to strong correlations (between 0.3576 to 0.5753). This excludes the worried about finances variable, which second period has a weak positive correlation (between 0.1232 to 0.177) except for the South states (medium positive correlation, 0.4765)) where the sliding window plots show a gradual negative slope rather than clear waves (Figure 6 (c)). The outcomes for the largest minimum and maximum correlation values for the second period was the same for all three variables, with the minimum being the northeast, and the maximum being the south.

Figure 7 and Table 2 shows the results of the eigenvector centrality values for all the mental health variables. The dynamic connectome plots using eigenvector centrality values shown in Figure 7 looks like the plots created by the correlation values in Figure 6. Although, Figures 7 (a) and (b) is less smooth, we see a similar shape and peak for the feeling anxious and feeling depressed variables. Following an initial decrease during the first period, both graphs show a sudden peak in the second period which plateaus and remains stable during the third period. This is a pattern also seen in Figure 6. Similarly, the worried about finances plot (Figure 7 (c)) also decreases sharply during the first period and does not fluctuate in the following periods, resembling the shape of Figure 6 (c).

From Table 2, we observed that during the first period for the feeling anxious variable, the northeast had the highest maximum value (0.7954) while the midwest had the smallest minimum eigenvector centrality value (0.1014). The minimum and maximum values during the second period were recorded in the northeast and South (0.1128 and 0.8252, respectively). This position flips during the third period, where the South now has the lowest minimum value (0.1344) and the Northeast has the largest maximum (0.5786).

For the dynamic connectome results of the feeling depressed variable, only two regions are recorded for the largest maximum and lowest minimum eigenvector centrality values. The midwest has the smallest minimum value for the first and second period (0.1446 and 0.1273, respectively) and the maximum value for the first period (0.8178). The second region was the northeast, seen with the largest maximum values in the second (0.7820) and third (0.6298) periods, as well as the minimum value (0.1622) in the third period.

The maximum and minimum regions for the first period of the worried about finances variable showed the south as the minimum (0.1617) and the midwest with the maximum (0.9493) for the eigenvector centrality results. In the second period, the south is recorded for both the lowest minimum and maximum values (0.0791 and 0.7831, respectively), and the midwest and west states had the minimum (0.1148) and maximum (0.6512) values for the third period, respectively.

Unlike the correlation values, there are not many similarities across the three variables’ eigenvector centrality values. We observed the south recorded for the maximum variable in the second period for both the feeling anxious and worried about finances variables. However, this is not reflected in feeling depressed. When looking at maximum and minimum eigenvector centrality values, the midwest and northeast show up the most frequently, especially for the feeling anxious and depressed variables. The south is seen for both the feeling anxious and worried about finances, while the West is only recorded once for worried about finances.

### Political dynamic connectome

The dynamic connectome for political party preferences of the states was conducted using sliding window analysis with snapshots of overlapping time windows of 30, resulting in correlation plots (see Figure 8) and eigenvector centrality values plots (see Figure 9). The maximum and minimum values for both data types are summarized in Tables 3 and 4.

In Figure 8 (a) there is a difference between Democratic and Republican states at around 30 to 80 days, where the correlation increases for Democratic states, but not as much for Republican states. It should also be mentioned that the correlation value at the start of the plot is higher in the democratic states compared to the Republican states. Like the regional dynamic connectome plots for demographic regions, Figure 8 (c) shows a negative slope with a lower plateau by the second period, while Figure 8 (a) and Figure 8 (b) start with a negative slope in the first period, have a peak in the second period and a lower plateau in the third.

Similar to the regional dynamic connectome of the geographic regions, the maximum values recorded during the third period are the lowest within the periods for all three variables. The minimum values are also extremely small numbers close to 0 (within -0.0111 to -0.0316), indicating negligible correlation. Across all three variables, the only consistent outcome was having the highest maximum correlation value in republican states during the second period.

From to Table 3, we observed that the lowest minimum and largest maximum correlation values (−0.0196 and 0.3994) for the first period in the feeling anxious variable was recorded for the Democratic states. During the second period, the minimum correlation value (−0.0316) remained in the Democratic states; the highest maximum correlation value (0.4394) was among the Republican states. In the third period, these positions are flipped, where the largest minimum value (−0.0181) was among the Republican states and the maximum within the Democratic states (0.0991).

For feeling depressed variable, the Republican states has the largest minimum correlation value across all three periods (−0.0277, -0.0293, and -0.0277). The largest maximum correlation for the first and third period are seen in the Democratic states (0.3365, and 0.0663), while the largest maximum correlation value was observed in the republican states for the second period (0.4507).

In the worried about finances variable, all minimum and maximum correlation values are largest among the Republican states. The largest minimum values for the three time period are (−0.0199, -0.0085, and -0.0077), and the largest maximum correlation values are (0.8104, 0.1735, and 0.1764).

Figure 9 and Table 4 shows the results of the eigenvector centrality values for all three mental health variables. Looking at the minimum and maximum values for the eigenvector centrality for the feeling anxious variable in Table 4 we observed that the Republican states had the minimum value (0.0804) during the first wave, while Democratic states had the maximum eigenvector centrality values (0.6633). In the second wave, the minimum and maximum values (0.1764 and 0.7153) were recorded for the Republican states. In the third wave, the minimum and maximum eigenvector centrality values (0.1810 and 0.4663) were seen within the Democratic states.

When observing the eigenvector centrality values for the feeling depressed variable, the lowest minimum and largest maximum values have the similar party outcome as the eigenvector centrality values for the feeling anxious variable; Republican states had the minimum values in the first and second period (0.1624 and 0.1173) and Democratic states (0.1176) in the third time period. A similar party outcome as feeling anxious was observed for the maximum eigenvector centrality values, namely Democratic states in the first and third period (0.6597 and 0.4621) and Republican in the second time period (0.7237).

In the political dynamic connectome results for worried about finances, all the lowest minimum and largest maximum eigenvector centrality values are among the Republican states. The lowest minimum values for the three time period are (0.1684, 0.1437 and 0.1490), and the largest maximum eigenvector centrality values are (0.9139, 0.5652 and 0.5483). Across the eigenvector centrality values for all three variables, the Republican states consistently recorded the highest maximum values during the second period.

## 4 Discussion

In this study, we applied clustering analysis from network theory to understand the connectivity and similarities between states and the impacts of COVID-19 on mental health across the country using COVID-19 related mental health indicators such as worried about finances, feeling depressed, and feeling anxious. We framed our results in terms of three time periods of three COVID-19 waves identified between March 2nd, 2021 to January 10th, 2022 using daily COVID-19 related cases, deaths, and hospitalizations (see Figure 1).

At the beginning of the pandemic several policies were put in place to curtail the outbreak; and several mandates were enacted to alleviate the public’s burden of the pandemic, see Section 2.1 for a list of some of the implemented policies and mandates. To understand the trends in people’s mental health during the pandemic we now investigate the possible effects of government policies and mandates from March 3rd, 2021 to January 10th, 2022. To assess the impact of these policies and mandates on people’s mental health, we evaluated how correlation values (from the regional and political dynamic connectomes) for each variable changed during the initial 30 days after a policy was implemented, and relate this change to the average reported percentages for the correlating mental health corresponding indicator. This allowed us to see if a higher correlation was indicative of an increasing or decreasing percentage of people experiencing the mental health indicators of interest. We used consensus clustering to group similar states together and dynamic connectome computed via sliding window analysis to create a time series of their relationship. The consensus clustering using the three mental health indicators shows correlation between the states. We discuss the outcome in detail below.

### Correlation in the south region

In the consensus clustering map shown in Figure 4, a noticeable clustering that resembled the south geographical region grouping shown in Figure 2 (a) can be observed for both the feeling anxious and worried about finances indicators. This similarity was also observed for the Republican party preference, as majority of the southern states are also Republican states, see Figure 2 (b).

We expected that the dynamic connectodome time series obtained from the sliding window analysis would lead to a high correlation within the states in the south. This is based on consensus clustering results showing many of the states within the south exhibiting similar mental health trends for both feelings of anxiety and worried about finances (see Figure 4). Thus, looking at the largest maximum and smallest minimum outcomes across the three periods (obtained from the three COVID-19 waves) for both geographical region and political party preference, we see that the south and Republican states consistently have the largest maximum value in the second period or wave between 100 and 200 days for the regional and political dynamic connectodome time series in Figures 6 and 8), respectively. This pattern is seen in both correlation and eigenvector centrality values when looking solely at the feeling anxious and worried about finances variable. Interestingly, only the consensus clustering map for feeling depressed lacked any distinct geographical or political clustering.

### Policy impact on feeling anxious and depressed

In Figures 6 and 8), we see that both feeling anxious and depressed variables have a similar wave pattern across the regional and political dynamic connectodome, they have several peaks in the first period, a single peak with high correlation during the second period, and a low plateau in the third. One difference between the two variables are the peaks around day 40 to 80 during the first period in average correlation especially for midwestern, northeastern (Figure 6 (a)), and Democratic states (Figure 8 (a)). The peaks for feeling anxious during this period is not as prominent as the peaks in the feeling depressed plots. These peaks shows an average 1.5% decrease in the midwest states reporting feelings of anxiety, a 2% decrease in the northeast, and a 2% decrease in Democratic states. Although the west also has these peaks, the correlation value is only weakly positive (0.21) compared to the midwest and the northeast (0.44 and 0.45, respectively) The presence of these peaks across the feeling anxious and feeling depressed variables during this time for the northeast and midwest may reflect the impact of policy changes on mental health indicators.

There were two policies introduced within 30 days prior to the start of these peaks seen between days 40 and 80. The first was a policy on April 19th (19th day mark) that guaranteed all adult Americans are eligible for COVID-19 vaccine, and the second a CDC guidance update on masking made on May 13, the 43^rd^ day mark. In their guidance, the CDC retracted their recommendation for fully vaccinated individuals to wear masks indoors or outdoors.

The introduction of a policy that would assist people in getting vaccinated and protecting themselves during the pandemic can reasonably be understood to work towards a decrease in reported percentages of anxiety and depression. The second policy however, is more ambiguous. While on one hand, individuals can consider this relaxing of guidance to indicate a decrease in severity and danger of the COVID-19 pandemic, others may see this as a premature decision and worry about the potential outcomes of making this announcement. Seeing how the states with an increased correlation all showed a decrease in feeling anxious and depressed indicators, it seems like these policies may have played a part in reducing anxiety and depression within the country, especially in the northeast and midwest regions. In addition to the policies, Figures 1 and 10 show how there had been an uptick in average COVID-19 cases, deaths and hospitalizations in democratic and northeastern states, which started to decrease around May to July of 2021. This timing aligned with the increase in political dynamic connectome correlation values for the feeling anxious and depressed variables, and a decrease in their corresponding indicator values. The decrease in COVID-19 related cases, deaths, and hospitalizations may have worked in synchrony with the policies mentioned above to alleviate people’s worries about COVID-19.

In addition to the peaks during the first period between days 40 to 80, the midwest also has another peak within this period for the feeling anxious connectome plots (Figure 6 (a)) with an initial weak correlation of 0.2. However, the feeling anxious indicator decreased in the first 12 days following the first window, resulting in increase in correlation value (medium positive correlation, 0.45). This increase is within 30 days of Biden signing a coronavirus relief bill, and it is possible that this government decision contributed to the decreasing feelings of anxiety among people in the midwest. A similar pattern is also seen for the feeling depressed indicator, where the correlation value increases over the first 14 days then sharply decreases. Like the feeling anxious variable, this fluctuation is within 30 days of the government decision, and is characterized by a decrease in the feeling depressed indicator across states within the midwest. This change in value tapers off, and is reflected by the decreasing correlation value on the graph.

In Figure 6 (b), the northeast also shows an increasing correlation in the feeling depressed variable over the first 21 days, reaching a medium positive correlation of 0.51 before decreasing. However, during the first 30 days (March 3rd to April 1st, 2021), the feeling depressed indicator increased, before slowly decreasing over the 21 days. The initial increase contradicts what we expected to see, as the introduction of the relief package should help alleviate financial and emotional stress. However, this lag in response may possibly be explained when considering that feelings of depression often take time to appear. Thus, it is understandable how the states in the northeast took longer to report a decrease in feelings of depression.

The most significant changes in the regional and political dynamic connectodome for feeling anxious and depressed variables happened during the second period, when regions and political parties peaked between the 110 and 165 day mark. During this time period, all regions and political parties experienced an increase in the feeling anxious and depressed indicators. Overall, the change in percentage was higher for feeling anxious compared to feeling depressed. Now compare this to the mental feeling anxious and depressed indicators, we see that the midwest had a 3% increase in reported feelings of anxiety, northeast had a 2.5% increase, west a 3% increase, and both Democratic and Republican states had 4% increase. The largest increase of 4.5% was seen in the southern states. For the percentage of individuals with feelings of depression, we see 1% increased in both midwest and the northeast, a 1.6% increase in the south, a 1.5% increase within the west, and a 1.7% increase among Democratic states. The highest increase of 1.8% was observed among Republican states. These increase in correlation begins roughly 15 days after the CDC announced that more than 50% of all COVID-19 cases were due to the Delta variant on July 3rd. While this announcement may not have directly resulted in the increase in value for feeling anxious and depressed indicators, we cannot over look how this announcement and correlation increase overlapped at essentially the same time as the increase we see in COVID-19 cases in Figures 1 and 10.

Keeping this CDC announcement in mind, we can conclude that the emergence of the Delta variant is a likely contributor to the sudden increase in feelings of anxiety and depression. As most nation-wide mandates had expired, it is likely that each state had to respond to the increase in COVID-19 cases individually, resulting in some states and regions being hit harder than others. In Figures 1 and 10 we see how the number of cases and deaths is much higher in the South and republican states. Assuming negative COVID-19 outcomes such as an increase in cases and deaths negatively impacts mental health, we would expect to see the hardest hit regions to also exhibit the largest increases in feeling anxious and depressed indicators. As expected, we found the south and the Republican states showed the largest percentage increase in mental health indicator values during the second period when the Delta variant began spreading.

### Policy impact on worries about finances

The regional and political dynamic connectome for the worried about finances variable decreased rapidly within the first 50 days, and maintained a lower plateau for the remainder of the periods, as seen in Figures 6 (c) and 8 (c). This is in contrast to the feeling anxious and feeling depressed variables which consistently showed a notable increase in correlation between 100 and 200 days in the dynamic connectome time series plot. The initial high correlation (all above 0.7 except for the northeast with 0.45) at day 0 for both geographic regional and political party preferences is indicative of how the first 30 days in the dynamic connectome showed the biggest collective trend among states. Looking at the reported values, the worried about finances indicator decreases rapidly across the regions and party preferences, during the first 30 days (from March 3rd to April 1st of 2021). Decreasing approximately 6% in the midwest, 3% for the northeast, 6% in the South, 5% in the West, 4% for Democratic states, and 6% for Republican states. The high correlation value reflects how all the states showed a decrease in the worried about finances indicator during the first few days of the data. Given the plateau following the initial negative slope, it can be assumed that the states went on to exhibit their own trends, and this interstate-level difference resulted in little to no correlation (0.2 and under, except for the south).

Following these decrease, the remaining data points for most states fluctuate within 2 to 3 percent at a slower rate post 30 day value. The initial decrease in the first period is the only change we see in the data, with the exception of the southern states that has a medium correlation peak (0.47) around the 110 - 135 day marks (Figure 6 (c)) showing an increase in the worried about finances indicator within the south. This uptick was also observed in the midwest and Republican states, albeit at a smaller increase, barely reaching a fairly weak positive correlation of 0.2.

The decrease in individuals’ worries about finances may be explained by the different policies relating to individual’s finances. As listed in Section 2.1, on March 11, 2021 Biden signed a coronavirus relief package which extended unemployment compensation and included a third stimulus check. However, while this event happened during the first 30 days of the collected survey responses, the data was already exhibiting a downward trend. It is likely that there are other external factors playing into this decrease; understanding how this policy may have impacted financial worries would require analysis on earlier time-points. Another possible explanation for the decreasing trend in financial worries may be an improvement in the employment rate. Compared with a peak civilian unemployment rate of 14.7% in April of 2020 ([27]), the unemployment rate was 6% during March and April of 2021, steadily decreasing to 4% by January of 2022, which marked the end of this study’s timeline. However, if all states had experienced a decreasing worry relating to finances, we should expect to see a higher correlation if worries continued to decrease.

Additional policies introduced later on such as a tax credit to decrease the burden of paid-leave on small businesses and non-profit organizations did not show any increase in correlation. It may be that by the end of the first period, rather than nation-wide policies, state-level government policies had more influence on the mental health of residents, and the direction each state took was not necessarily related to or dictated based on geographical regions or political party preference. We hypothesized that policies impacting a large number of people, such as eviction moratoriums and unemployment benefits ending would result in an increase in correlation values across all groups and states, however the average correlations in both the second and third period remain stable 30 days after these policies are implemented.

Another possibility arises when considering the slight peak shown by the south during the second period. The 110 day mark (July 20th, 2021) is only 13 days after the CDC announced the COVID-19 Delta variant as responsible for over 50% of all COVID-19 cases. In the same line, we see a larger spike in COVID-19 cases for both the south and Republican states in late July (Figures 1 and 10). It could be possible that states most heavily impacted with COVID-19 took similar methods to address the Delta variant, resulting in a brief increase in correlation among the south.

### Comparison with Fulk et. al [10] results

Fulk et. al [10] investigated the effect that several variables related to the COVID-19 pandemic had on anxiety and depression in the general population during the same time period as this study. They presented their findings in a bidirectional format and found that variables directly related to COVID-19 (i.e. COVID-19 incidence and death) had a significant positive effect on anxiety and depression in several states. In our study, we found that the correlation in anxiety and depression between states increased dramatically in the second time period. This coincided with the rise of the Delta variant of SARS-CoV-2, which had significantly increased transmissibility compared to previously dominant strains of SARS-CoV-2. One advantage of using sliding window and clustering analyses over the bidirectional approach taken in [10] is that we are able to see greater variability through time and across the country. Figures 1, 2, 5, and 6 in [10] show that an increase in the incidence of COVID-19 led to an increase in the percentage of individuals that reported feeling anxious/depressed in a large number of states. We can make similar conclusions based on the results in Figure 3 (a) and in conjunction with Figure 4 (a) and (b). Further, we can see which states were behaving similarly (e.g. the south cluster) in their correlation over the entire period of the analysis and we don’t have to rely solely on significant results to draw our conclusions. An additional insight that the results of [10] pertains to why certain states are commonly clustered together. For example, Florida and Texas were significant in several of the analyses of the effect of COVID-19 incidence on anxiety and depression, which may indicate that their populations were responding similarly to the growing pandemic and thus explains why they are grouped together with respect to each of the variables evaluated in our study. The variables related to social isolation in [10] were less informative to our study and seemed to vary significantly more compared to variables related to COVID-This may indicate that the trends seen in our indicators were impacted more by COVID-19 related issues than social isolation.

**Figure 3:**
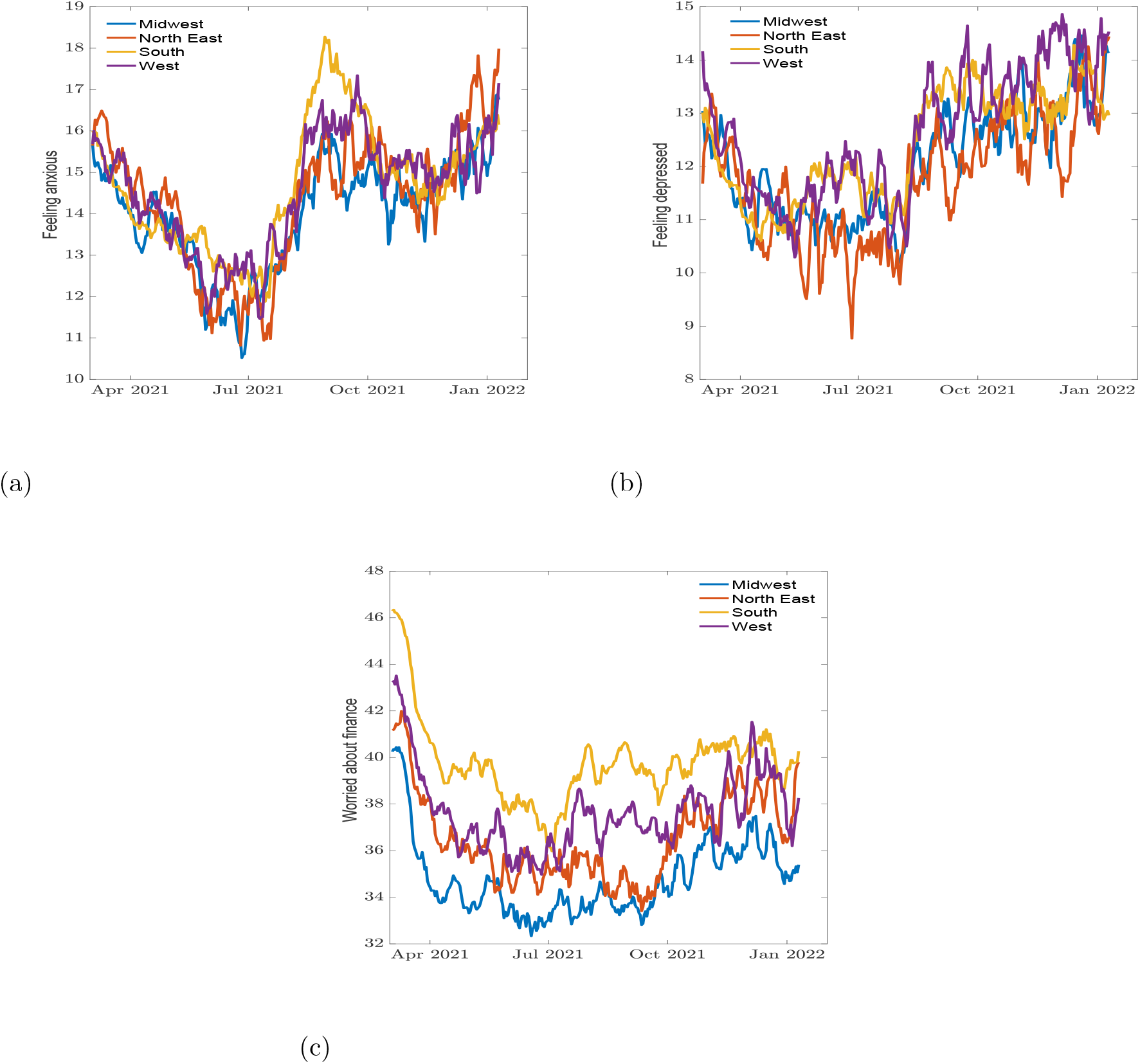
Percentage of individuals (a) feeling anxious, (b) feeling depressed, and (c) worried about finances across demographic regions.

**Figure 4:**
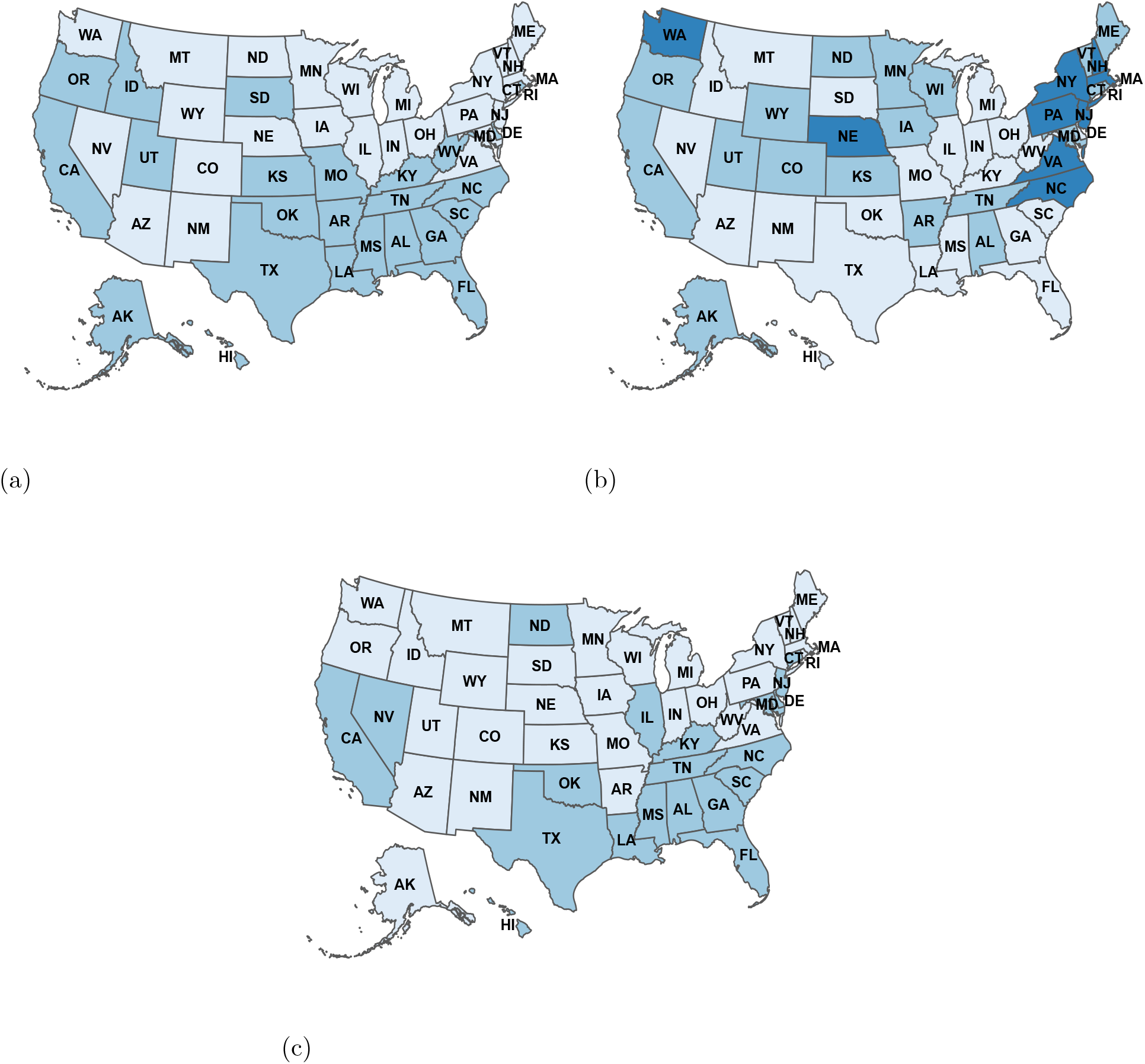
Country-wide maps grouping states with similar trends between 3/2/2021 to 1/10/2022 for (a) feeling anxious, (b) feeling depressed and (c) worried about finances indicators. The states with light blue, blue, and dark blue are in the same cluster.

**Figure 5:**
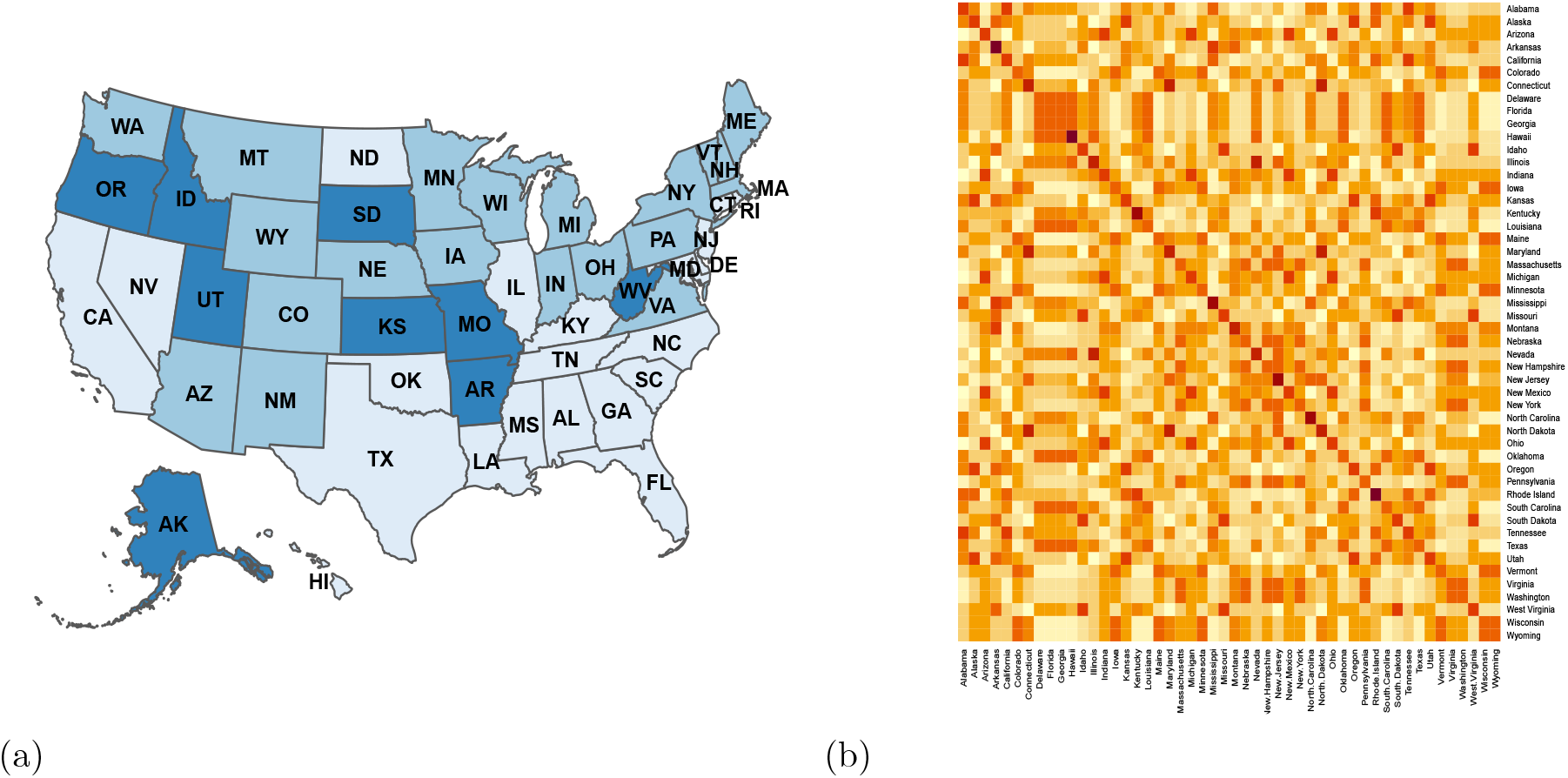
Allegiance clustering for feeling anxious, feeling depressed, and worried about finances indicators. (a) Allegiance map; (b) Allegiance Heat-map. The states with the same color (light blue, blue, or dark blue) are in the same cluster.

**Figure 6:**
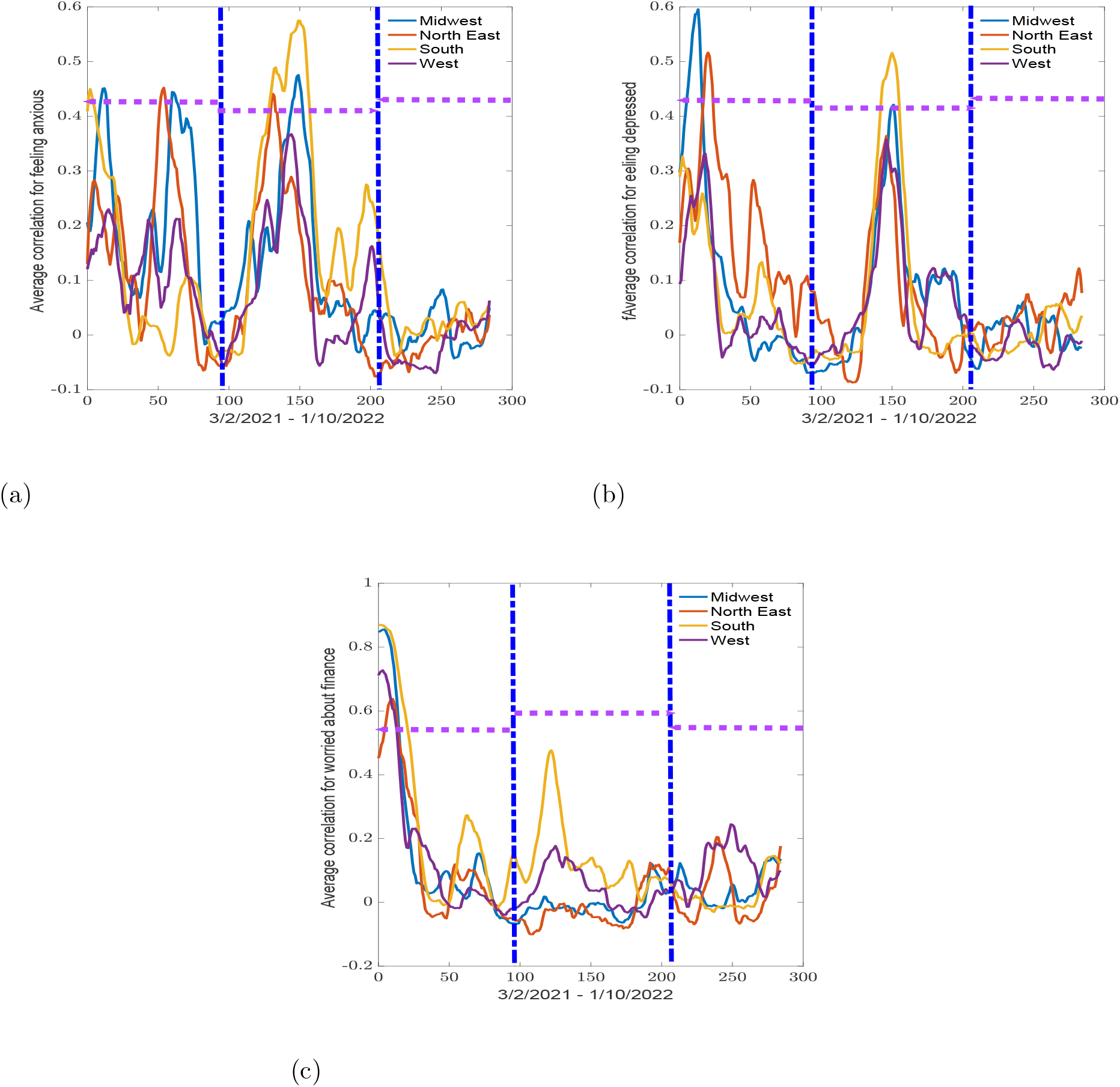
Average correlation for states in Midwest, Northeast, South, and West regions between 4/3/2021 to 1/10/2022 for(a) feeling anxious, (b) feeling depressed, and (c) worried about finances variables. The vertical blue dot-dash lines indicate the start and end of the epidemic waves observed in Figure 1 and the horizontal dash lines indicate the width of the waves.

**Figure 7:**
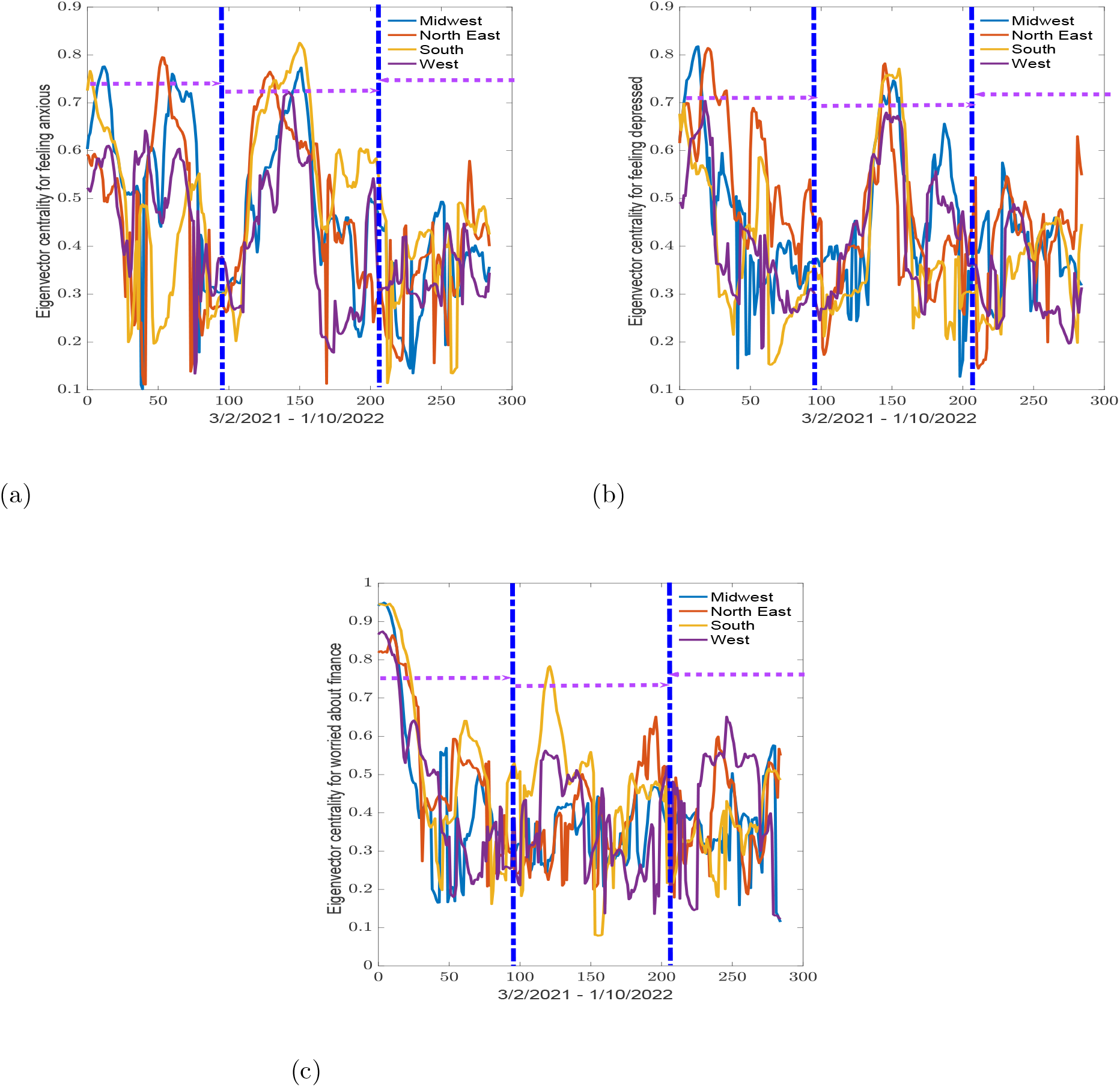
Average eigenvector centrality for states in midwest, northeast, south, and west regions between 4/3/2021 to 1/10/2022 for (a) feeling anxious, (b) feeling depressed, and (c) worried about finances variables. The vertical blue dot-dash lines indicate the start and end of the epidemic waves observed in Figure 1 and the horizontal dash lines indicate the width of the waves.

**Figure 8:**
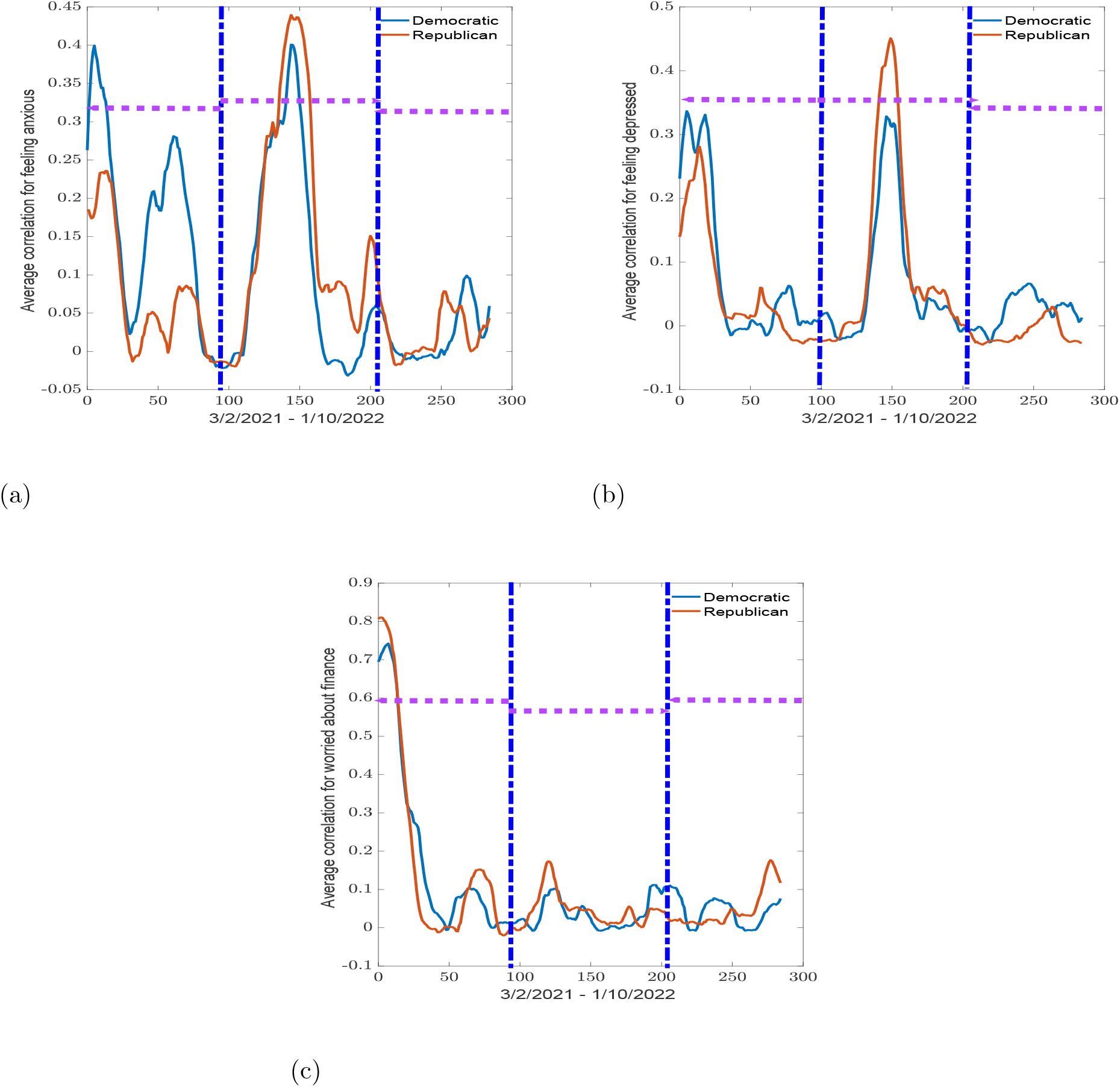
Average correlation within Democratic and Republican states between 4/3/2021 to 1/10/2022 for (a) feeling anxious, (b) feeling depressed, and (c) worried about finances variables. The vertical blue dot-dash lines indicate the start and end of the epidemic waves observed in Figure 1 and the horizontal dash lines indicate the width of the waves.

**Figure 9:**
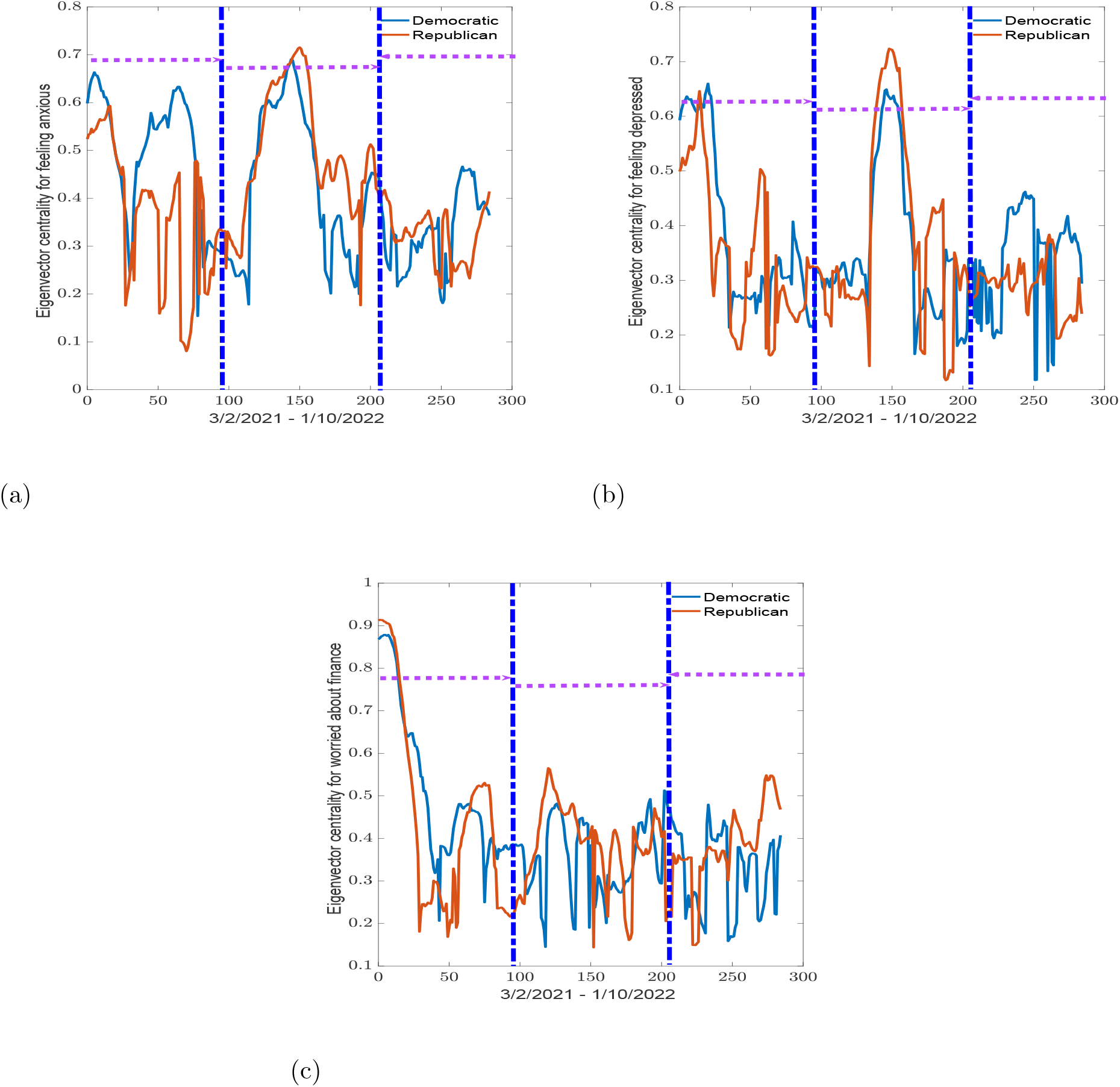
Average eigenvalue centrality for Democratic and Republican states between 4/3/2021 to 1/10/2022 for (a) feeling anxious, (b) feeling depressed, and (c) worried about finances variables. The vertical blue dot-dash lines indicate the start and end of the epidemic waves observed in Figure 1 and the horizontal dash lines indicate the width of the waves.

**Figure 10:**
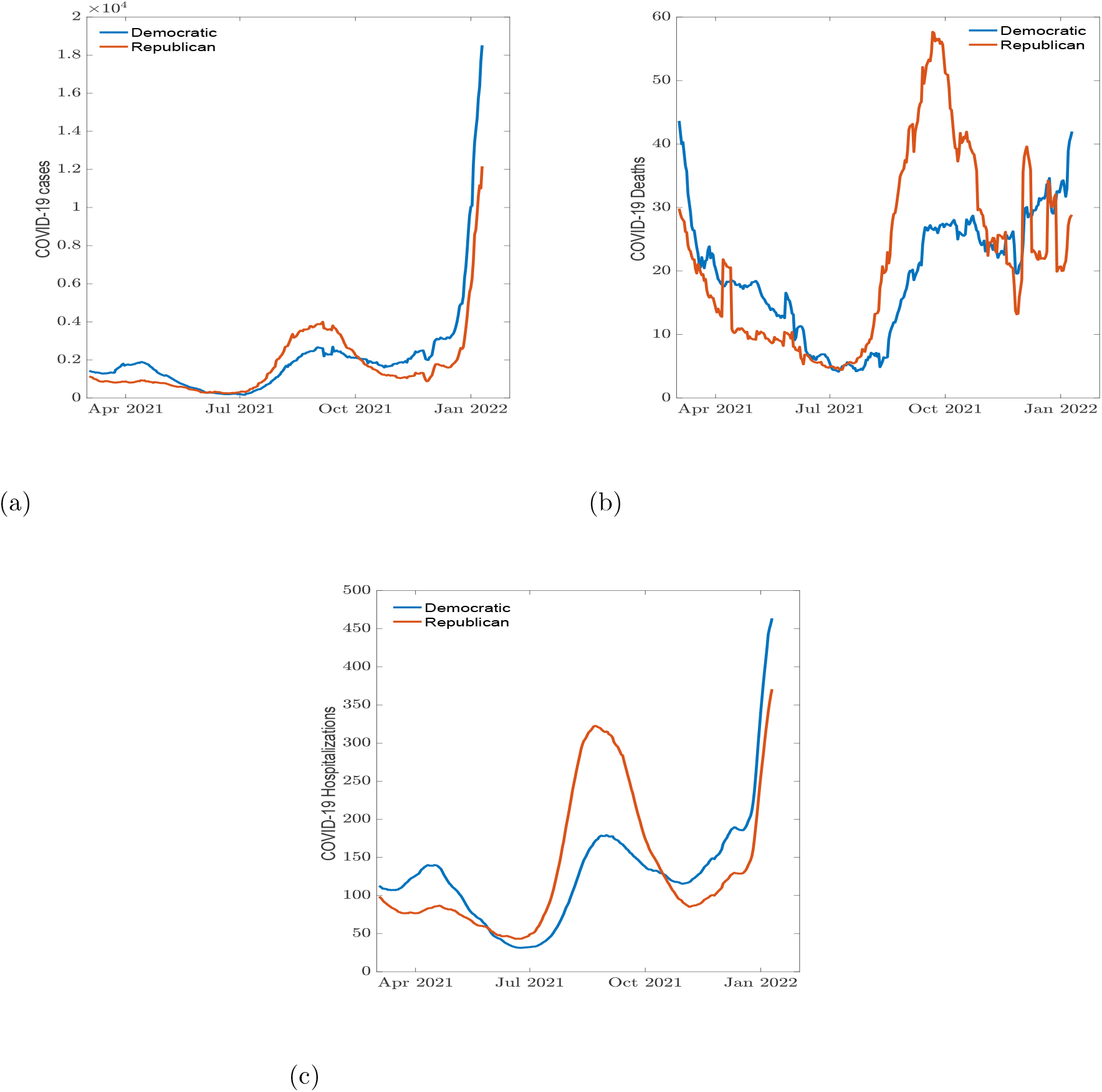
Daily COVID-19 (a) cases, (b) reported deaths, and (c) hospitalization cases between state political preference.

**Figure 11:**
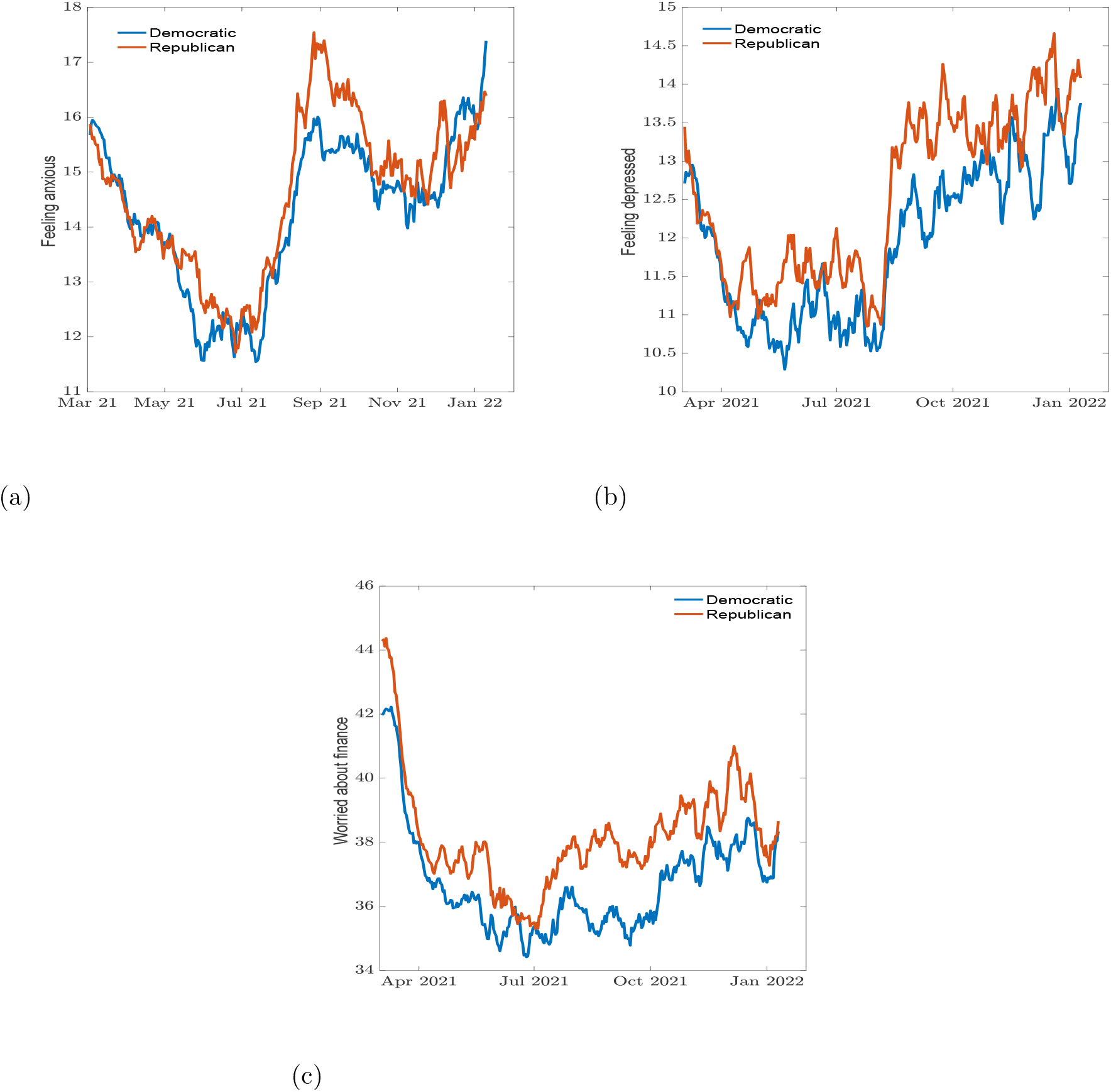
Percentage of individuals (a) feeling anxious, (b) feeling depressed, and (c) worried about finances between state political preference.

## Conclusion

Numerous studies have observed an increase in negative mental health conditions across the United States in parallel with the emergence of COVID-19 [9, 10, 12, 21]. Increase in COVID-19 incidence and deaths has been implicated in increased feelings of anxiety and depression across multiple states [10]. In this work, we applied network and clustering analysis to study the connectivity and similarities between states and how COVID-19 impacts mental health across the country using the “mental health functional connectome” obtained from the covariance matrix of mental health indicators, namely worried about finances, feeling anxious and feeling depressed. We summarize our findings below:

i. The southern states and Republican party showed similar trends for feeling anxious and worried about finances indicators between March 3rd, 2021 and January 10th, 2022.
ii. The mental health indicator for feeling depressed showed no identifiable communities resembling geographical regions or political party affiliation.
iii. The highest correlation values for the feeling anxious and feeling depressed variable seemingly overlapping with an increase in COVID-19 related cases, deaths, hospitalizations, and rapid spread of the COVID-19 Delta variant.

### Future steps and limitations

Results from this study showed a decrease in worries about finances variable across the country in the correlation among states and political affiliation; conducting further research using more survey data, particularly from dates earlier than March 3rd, 2021 may provide further insight into possible reasons for the decrease.

Anxiety can be fueled by sources such as media exposure and negative experiences with COVID-19; however, the government and state response to increasing cases may also alleviate or exacerbate feelings of anxiety. Since the data collected for this time-frame began in the early half of 2021, most lockdown measures had ended, and organizations such as the CDC were consistently issuing and updating health mandates and guidance information. In addition to national government responses, some states were actively loosening restrictions compared to others that maintained some level of safety measures to combat the appearance of COVID-19 variants. Whether participants felt safe or worry in response to loosening restrictions in that state could influence how they responded on the survey, affecting how a state related to other regions, and are a potential area for future research. Previous studies have identified certain protective factors against symptoms of anxiety, including old age, higher education, being male, and a good economic situation [15]. Comparing these factors with the map distribution for anxiety, such as the average level of education attained in each state, could provide more insight as well.

Given how close certain policies were implemented, such as the expiration of an eviction moratorium as well as unemployment benefits happening within 11 days of each other, it is difficult to speculate how a given policy may have impacted a mental health indicator. It also fully possible that the impact a policy could have on a variable - if any - arose due to the anticipation of a policy being introduced or a mandate being lifted rather than after it is implemented. Since we observed any correlation fluctuation 30 days after a policy was introduced rather than before, we have not taken anticipatory effects into consideration. In addition, by using a 30 day window, our sliding window analysis may have become less sensitive to short-term changes in correlation, as the values would need to collectively increase or decrease for a longer span of time in order to be picked up by the analysis.

Sliding window analysis requires the researchers to balance parameters such as window size prior to conducting the analysis. However, as there is no established standard for setting a window size, it is up to each researcher to choose a parameter that best fits their research question and intentions resulting in ambiguity. Incorporating different methods of centrality and network analysis would allow for more insight, stability and interpretation on the changes in values over time, which the sliding window analysis and eigenvector centrality values may not fully provide.

Another limitation observed in this study is the difficulty of interpreting the feelings of depression variable. This mental health indicator is difficult to measure due to the long-term nature of depression, as well as the overlapping effects certain policy changes may have on the mental health indicators of interest. Unlike anxiety and worries about finances, depressive symptoms may take much longer to manifest themselves in any person, making it difficult to determine what policies impacted this indicator. Any singular policy change may also affect a multitude of mental health indicators, and the extent of which each indicator is affected cannot readily be analyzed.

## Data Availability

Data used are available at the link https://www.pnas.org/doi/full/10.1073/pnas.2111454118.
All code, data, and results are available upon request from the corresponding author.

## Data Availability

Data used are available at the link https://doi.org/10.1073/pnas.2111454118

All code, data, and results are available upon request from the corresponding author.

## Author contributions

Hiroko Kobayahsi: formal analysis, writing - original draft preparation, reviewing and editing; Raul Saenz-Escarcega: Writing - original draft preparation, reviewing and editing, Alexander Fulk: Writing - formal analysis, original draft preparation, eviewing and editing, and Folashade B. Agusto: Conceptualization, project administration, supervision, writing-reviewing and editing, funding acquisition.

## Acknowledgement

This research was funded by the National Science Foundation, grant number DMS2028297 and DMS2230117.

## A Political: Cases, deaths, and hospitalization data

The daily COVID-19 related cases, deaths, and hospitalizations were grouped based on each state’s political siding showing waves in the spread of COVID-19.

## B Political: Mental health data

The survey results on the mental health indicators based on political preference.

## Footnotes

2 Policy guideline that affects finances

3 Policy guideline that affects feeling anxious and depressed

## References

[1] Abdul Basit Adeel, Michael Catalano, Olivia Catalano, Grant Gibson, Ezgi Muftuoglu, Tara Riggs, Mehmet Halit Sezgin, Olga Shvetsova, Naveed Tahir, Julie VanDusky-Allen, et al. Covid-19 policy response and the rise of the sub-national governments. Canadian Public Policy, 46(4):565–584, 2020.

[2] Lara B Aknin, Jan-Emmanuel De Neve, Elizabeth W Dunn, Daisy E Fancourt, Elkhonon Goldberg, John F Helliwell, Sarah P Jones, Elie Karam, Richard Layard, Sonja Lyubomirsky, et al. Mental health during the first year of the covid-19 pandemic: A review and recommendations for moving forward. Perspectives on psychological science, 17(4):915–936, 2022.

[3] American Medical Association (AMA). The role of data collection in the COVID-19 pandemic. https://www.ama-assn.org/delivering-care/health-equity/role-data-collection-covid-19-pandemic. Accessed: 2022-07-27.

[4] Enrico Amico and Iulia Martina Bulai. How political choices shaped covid connectivity: The italian case study. Plos One, 16(12):e0261041, 2021.

[5] Blazej M Baczkowski, Tom Johnstone, Henrik Walter, Susanne Erk, and Ilya M Veer. Sliding-window analysis tracks fluctuations in amygdala functional connectivity associated with physiological arousal and vigilance during fear conditioning. Neuroimage, 153:168–178, 2017.

[6] Center for Systems Science and Engineering (CSSE). COVID-19 Data Repository at Johns Hopkins University. https://github.com/CSSEGISandData/COVID-19. Accessed: 2021-06-15.

[7] Rastko Ciric, Jason S Nomi, Lucina Q Uddin, and Ajay B Satpute. Contextual connectivity: A framework for understanding the intrinsic dynamic architecture of large-scale functional brain networks. Scientific reports, 7(1):1– 16, 2017.

[8] Domenico Cucinotta and Maurizio Vanelli. Who declares covid-19 a pandemic. Acta Bio Medica: Atenei Parmensis, 91(1):157, 2020.

[9] Mark É Czeisler, Rashon I Lane, Emiko Petrosky, Joshua F Wiley, Aleta Christensen, Rashid Njai, Matthew D Weaver, Rebecca Robbins, Elise R Facer-Childs, Laura K Barger, et al. Mental health, substance use, and suicidal ideation during the covid-19 pandemic—united states, june 24–30, 2020. Morbidity and Mortality Weekly Report, 69(32):1049, 2020.

[10] Alexander Fulk, Raul Saenz-Escarcega, Hiroko Kobayashi, Innocent Maposa, and Folashade Agusto. Assessing the impacts of covid-19 and social isolation on mental health in the united states of america. medRxiv, 2022.

[11] Matthew L Goldman, Benjamin G Druss, Marcela Horvitz-Lennon, Grayson S Norquist, Kristin Kroeger Ptakowski, Amy Brinkley, Miranda Greiner, Heath Hayes, Brian Hepburn, Shea Jorgensen, et al. Mental health policy in the era of covid-19. Psychiatric Services, 71(11):1158–1162, 2020.

[12] David Gunnell, Louis Appleby, Ella Arensman, Keith Hawton, Ann John, Nav Kapur, Murad Khan, Rory C O’Connor, Jane Pirkis, Eric D Caine, et al. Suicide risk and prevention during the covid-19 pandemic. The Lancet Psychiatry, 7(6):468–471, 2020.

[13] Thomas Hale, Anna Petherick, Jessica Anania, Bernardo Andretti, Noam Angrist, Roy Barnes, Thomas Boby, Emily Cameron-Blake, Alice Cavalieri, Martina Di Folco, Benjamin Edwards, Lucy Ellen, Jodie Elms, Rodrigo Furst, Liz Gomes Ribeiro, Kaitlyn Green, Rafael Goldszmidt, Laura Hallas, Beatriz Kira, Maria Luciano, Saptarshi Majumdar, Thayslene Marques Oliveira, Radhika Nagesh, Toby Phillips, Annalena Pott, Julia Sampaio, Helen Tatlow, Adam Wade, Samuel Webster, Andrew Wood, Hao Zha, and Yuxi Zhang. Variation in Government Responses to COVID-19, Version 13.0. http://www.bsg.ox.ac.uk/covidtracker, 2022. Blavatnik School of Government Working Paper.

[14] Laura Hallas and Saptarshi Majumdar Monika Pyarali Andrew Wood Thomas Hale Ariq Hatibie, Rachelle Koch. Variation in US states’ COVID-19 policy responses. https://www.bsg.ox.ac.uk/research/publications/variation-us-states-responses-covid-19, 2021.

[15] Angela M Kunzler, Nikolaus Röthke, Lukas Günthner, Jutta Stoffers-Winterling, Oliver Tüscher, Michaela Coenen, Eva Rehfuess, Guido Schwarzer, Harald Binder, Christine Schmucker, et al. Mental burden and its risk and protective factors during the early phase of the sars-cov-2 pandemic: systematic review and meta-analyses. Globalization and health, 17(1):1–29, 2021.

[16] Joseph Lee Rodgers and W Alan Nicewander. Thirteen ways to look at the correlation coefficient. The American Statistician, 42(1):59–66, 1988.

[17] Mark Abadi. Even the US government can’t agree on how to divide up the states into regions. https://www.businessinsider.com/regions-of-united-states-2018-5. Accessed: 2022-08-24.

[18] Emma E McGinty, Rachel Presskreischer, Hahrie Han, and Colleen L Barry. Psychological distress and loneliness reported by us adults in 2018 and april 2020. Jama, 324(1):93–94, 2020.

[19] Stefano Monti, Pablo Tamayo, Jill Mesirov, and Todd Golub. Consensus clustering: a resampling-based method for class discovery and visualization of gene expression microarray data. Machine learning, 52(1):91–118, 2003.

[20] National Geographic. United States Regions. https://education.nationalgeographic.org/resource/united-states-regions. Accessed: 2022-07-27.

[21] Nirmita Panchal, Rabah Kamal, Kendal Orgera, Cynthia Cox, Rachel Garfield, Liz Hamel, and Priya Chidambaram. The implications of covid-19 for mental health and substance use. Kaiser family foundation, 21, 2020.

[22] Elise Paul, Andrew Steptoe, and Daisy Fancourt. Attitudes towards vaccines and intention to vaccinate against covid-19: Implications for public health communications. The Lancet Regional Health-Europe, 1:100012, 2021.

[23] Francisco Perez-Arce, Marco Angrisani, Daniel Bennett, Jill Darling, Arie Kapteyn, and Kyla Thomas. Covid-19 vaccines and mental distress. PLoS One, 16(9):e0256406, 2021.

[24] Joshua A Salomon, Alex Reinhart, Alyssa Bilinski, Eu Jing Chua, Wichada La Motte-Kerr, Minttu M Rönn, Marissa B Reitsma, Katherine A Morris, Sarah LaRocca, Tamer H Farag, et al. The us covid-19 trends and impact survey: Continuous real-time measurement of covid-19 symptoms, risks, protective behaviors, testing, and vaccination. Proceedings of the National Academy of Sciences, 118(51):e2111454118, 2021. Accessed: 2021-06-15.

[25] Vincent A. Traag, Ludo Waltman, and Nees Jan Van Eck. From louvain to leiden: guaranteeing well-connected communities. Scientific reports, 9(1):1– 12, 2019.

[26] Megh M Trivedi and Anirudha Das. Did the timing of state mandated lockdown affect the spread of covid-19 infection? a county-level ecological study in the united states. Journal of Preventive Medicine and Public Health, 54(4):238, 2021.

[27] U.S. Bureau of Labor Statistics. Civilian unemployment rate. https://www.bls.gov/charts/employment-situation/civilian-unemployment-rate.htm. Accessed: 2022-09-12.

[28] Vincent A. Traag. leidenalg. https://github.com/vtraag, 2020. Accessed: 2022-08-23.

[29] Yunhe Wang, L. Shi, Jianyu Que, Qingdong Lu, Lin Liu, Zhengan Lu, Yingying Xu, Jiajia Liu, Yankun Sun, Shiqiu Meng, et al. The impact of quarantine on mental health status among general population in china during the covid-19 pandemic. Molecular psychiatry, 26(9):4813–4822, 2021.

[30] Wikipedia. Political party strength in U.S. states. https://en.wikipedia.org/wiki/Political_party_strength_in_U.S._states. Accessed: 2022-09-07.

[31] World Health Organization (WHO). COVID-19 pandemic triggers 25 percent increase in prevalence of anxiety and depression worldwide. https://www.who.int/news/item/02-03-2022-covid-19-pandemic-triggers-25-increase-in-prevalence-of-anxiety-and-depression-worldwide. Accessed: 2022-07-27.

[32] World Health Organization (WHO). The impact of COVID-19 on mental health cannot be made light of. https://www.who.int/news-room/feature-stories/detail/. Accessed: 2022-07-27.

[33] World Health Organization (WHO). WHO Coronavirus (COVID-19) Dash-board. https://covid19.who.int/. Accessed: 2022-07-27.

